# A simple model for how the risk of pandemics from different virus families depends on viral and human traits

**DOI:** 10.1101/2021.03.17.21253813

**Authors:** Julia Doelger, Arup K. Chakraborty, Mehran Kardar

## Abstract

Different virus families, like influenza or corona viruses, exhibit characteristic traits such as typical modes of transmission and replication as well as specific animal reservoirs in which each family of viruses circulate. These traits of genetically related groups of viruses influence how easily an animal virus can adapt to infect humans, how well novel human variants can spread in the population, and the risk of causing a global pandemic. Relating the traits of virus families to their risk of causing future pandemics, and identification of the key time scales within which public health interventions can control the spread of a new virus that could cause a pandemic, are obviously significant. We address these issues using a minimal model whose parameters are related to characteristic traits of different virus families. A key trait of viruses that “spillover” from animal reservoirs to infect humans is their ability to propagate infection through the human population (fitness). We find that the risk of pandemics emerging from virus families characterized by a wide distribution of the fitness of spillover strains is much higher than if such strains were characterized by narrow fitness distributions around the same mean. The dependences of the risk of a pandemic on various model parameters exhibit inflection points. We find that these inflection points define informative thresholds. For example, the inflection point in variation of pandemic risk with time after the spillover represents a threshold time beyond which global interventions would likely be too late to prevent a pandemic.

## 1. Introduction

Infectious disease pandemics and epidemics caused by newly emergent pathogens can lead to major health and economic crises and have been on the rise in recent decades [1, 2, 3]. We, the inhabitants of the twenty-first century, have been cruelly reminded of the negative impacts of a global pandemic by the ongoing COVID-19 crisis [4, 5].

Major infectious disease epidemics and pandemics are often the result of pathogens that usually circulate in animals becoming capable of infecting humans [2]. Such jumps of pathogens from animals to humans are called zoonotic shifts. For viruses, especially those with RNA genome, such events happen when an animal virus adapts to human hosts, sometimes via an intermediate animal. Such adaptation can occur via mutation and/or re-assortment or recombination [6, 7]. RNA viruses mutate continuously, and recombination or re-assortment processes can happen when two or more viral strains infect the same cell in an animal or a human. If a virus strain that emerges in this fashion can infect humans, is reasonably contagious and reasonably lethal, frightening pandemics can occur. This is because few, if any, humans have immunity for this new pathogen, which can lead to a rapid spread of the disease [8], especially in our modern densely populated and connected world. If a not insignificant fraction of infected people require hospitalization, the medical system can be overwhelmed, and deaths and lockdowns exact an enormous human and economic toll.

For effective prevention and control of pandemics it would be helpful if one could anticipate future disease outbreaks. Various methods for prediction of the severity of disease outbreaks have been used [9, 10], but they usually focus on viruses that already circulate in humans, and therefore, the predictions tend to be rather specific and often short-term. Predicting the emergence of zoonotic strains more generally, however, presents major difficulties [11]. These novel strains are created from complex and often random interactions between humans, animals and their respective pathogens, via many mutations and recombinations/re-assortments and this makes the specific evolutionary and transmission paths leading to successful human strains hard to predict [6, 7, 12, 13]. Additionally there is a lack of data on the spectrum of circulating animal viruses as well as on their potential to cause human disease [12, 14]. Finally, most newly emerging viruses do not cause major outbreaks in humans. Pandemics are thus rare events, making their prediction from data especially difficult [15].

Even if precise predictions cannot be made regarding the viruses that are most likely to cause pandemics, and under what circumstances, it is important to obtain some mechanistic understanding of the phenomena that underlie answers to these questions. Specifically, can we obtain mechanistic insights into the relative risk of pandemics emerging due to viruses from different families that have different traits? Genetically related virus families evolve subject to a similar fitness landscape whose topology is determined by core viral traits such as genome length, typical mode of infection, replication and transmission. The traits of the animal host species in which the virus family circulates and their similarity and contact with humans are also important [16]. If we can describe a virus family with common traits and trait distributions as a unit, we can learn about their relative risk of causing pandemics, and the mechanism underlying these risk factors without making assumptions about specific evolutionary processes.

Here, we develop a pandemic risk model and employ it to calculate the probability and frequency for a pandemic to emerge from virus families with different characteristics. These characteristics include: the fitness distribution and zoonotic shift frequency of distinct viral types that circulate in animal hosts; the characteristics of the human host population in relation to the virus, which influence the growth and transmission of the disease from a local outbreak to the global human population. Human traits/behavior in particular influence the nature of the temporal variation of disease growth rate after initial emergence.

## 2. Description of the pandemic risk model

A sketch of the key features of our model is shown in Fig. 1. New functional human variants from a specific viral type are assumed to emerge at a zoonotic shift rate *µ*_shift_. Functional here means that these viruses are able to infect and reproduce in humans and have a positive human-to-human transmission rate. But the rate of replication and transmission can be very small for poorly adapted variants and the effective disease growth rate, given infection and recovery rates, can even be negative. Each such novel human strain is created by a complex interaction between animal and human hosts and between their respective, related viruses. This can also include intermediate hosts that form a link between the reservoir host and humans. In general, the process can involve various jumps between different host species and multiple mutation and recombination/re-assortment events. The zoonotic shift rate *µ*_shift_ therefore depends on various viral and host factors [16, 17]. The genetic similarity between animal reservoir hosts and humans determines how much an animal virus typically needs to change to adapt to the human host environment [15]. Thus we expect that animal hosts that are more similar to humans produce more frequent adaptations leading to a higher *µ*_shift_ than less similar animals. Physical proximity, including the extent and nature of interactions between reservoir and intermediate animal hosts and humans is another important factor influencing the zoonotic shift rate, since it determines the frequency, at which viral exchanges between the species occur [15]. Wild birds, for example, form a reservoir for many subtypes of influenza A virus, and most spillovers to humans have happened via the infection of humans by livestock hosts such as poultry and pigs, which are in close contact with humans [18].

**Figure 1.**
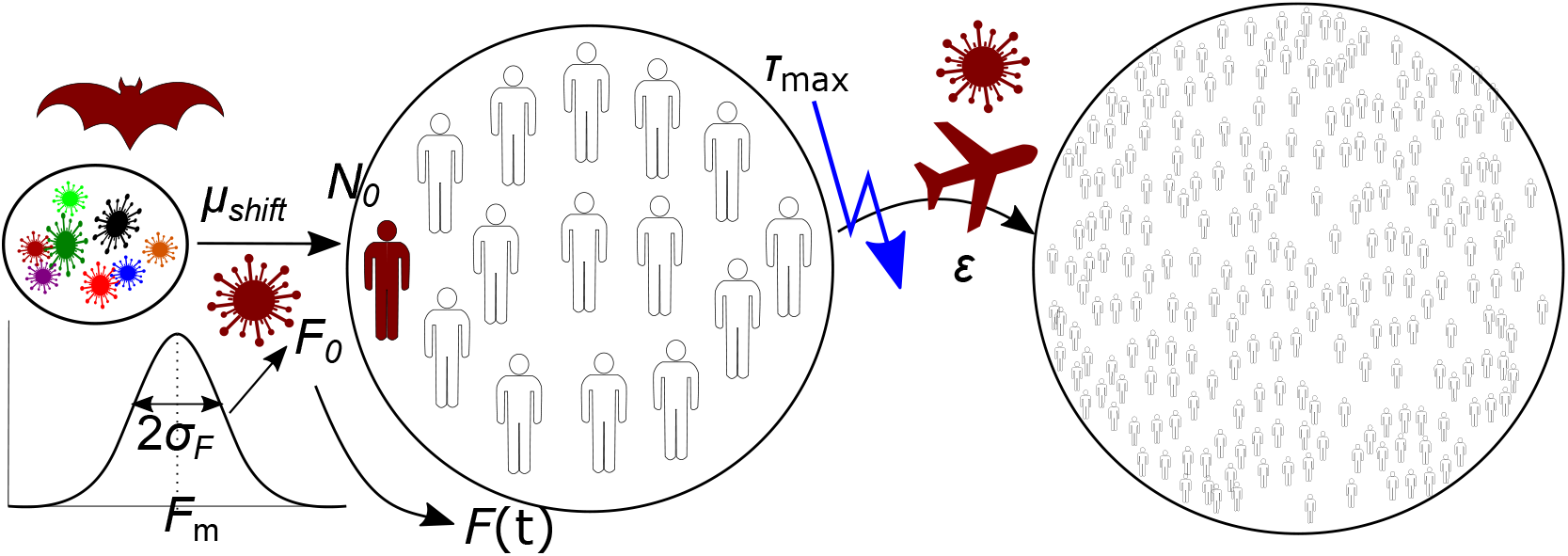
Schematic depiction of the model framework: A viral type, here shown as a group of viruses of different colors on the left, produces human variants by zoonotic shift (a bat is an example animal origin). Human functional variants, with initial fitness *F*_0_ drawn from a fitness distribution with mean *F*_m_ and variance 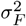 that characterizes their type, emerge at rate *µ*_shift_ and initially infect *N*_0_ people in the infection center (left circle with humans, colored white if susceptible, red if infected). The disease spreads in the infection center with effective rate *F* (*t*), which may change with time. Transmission to the rest of the world (right circle with many humans) that triggers a pandemic, happens at rate *E*, until a time *τ*_max_. This is the time before which the emergence and spread of a new disease is not noticed internationally, and so no restrictions to travel are put in place until then.

Each new human variant, being created at rate *µ*_shift_, is then represented with a fitness *F*_0_, which is the initial effective growth rate of the disease in the human population. The fitness parameter *F*_0_ is non-dimensional and represents infection rate minus recovery rate, times the typical infection period. This relates to the (basic) reproduction number *R* = *F*_0_ + 1, a standard parameter used in epidemiological models [10], which describes the mean number of secondary infections caused by one infected person.

In our model, fitness is drawn from a normal distribution *f*_*F*_ (*F*_0_| *F*_m_, *σ*_*F*_) with mean *F*_m_ and variance 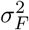, which represent the traits of a specific viral type evolving on a complex fitness landscape to become capable of infecting and potentially thriving in a human population. We scale all times and rates in our model with the typical infection period, which is equal to the time it takes for a person to recover from infection. The recovery rate in non-dimensional form is thus equal to 1.

The fitness depends on both the infectivity of the virus and on host characteristics such as population density and social interaction networks. We regard *F*_m_ as a typical initial fitness of novel functional variants of a particular type spreading in a human population without human intervention. *F*_m_ can also depend on the genetic similarity of humans and the reservoir hosts because viruses that are well adapted to a very distant reservoir host will require many changes to be well adapted to humans. Since such dramatic changes are unlikely, *F*_m_ would be small. The variance 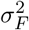 expresses variations of viral traits that lead to different rates of spread for different functional variants of the same type.

In the population where the disease first emerges, which we call the infection center, we assume that a small number *N*_0_ of initial human hosts are directly infected by animal contact. On average the outbreak, i.e. the mean number *N* of infections, grows as

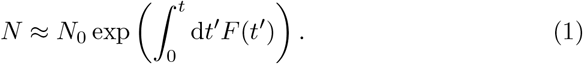

Fitness (starting from *F* (*t* = 0) = *F*_0_) generally varies with time, for example due to human interventions, environmental fluctuations, or reduction of the number of susceptible individuals with increasing population immunity.

During the spread of the disease in the infection center, individuals can travel away from the original center and transmit the disease to the rest of the world. We assume that each infected person in the infection center can seed a global outbreak at a rate *E*, and that a pandemic is caused when one such seed has emerged, which can then quickly spread globally. The “world transmission rate” *E* includes a measure of connectedness of the infection center with the rest of the world and transmissibility of the virus. The parameter *τ*_max_ depends on the rapidity with which the emergence of a new virus is understood. *τ*_max_ is large for a virus that causes a disease like COVID-19 with many asymptomatic yet infectious cases and a relatively low death rate, and smaller for the opposite disease characteristics. For example, the Ebola, Zika, and MERS epidemics were characterized by a smaller *τ*_max_ compared to COVID-19. This is because people infected with Zika, Ebola, or MERS quickly became very ill and a large fraction died [19], thus alerting the world to a deadly virus before it had time to spread widely. Diseases in this category are comparatively easily detected and subsequently controlled at the location of the outbreak, before significant international transmissions.

Since each infected person can seed a pandemic with rate *ϵ*, the total rate of pandemic seeding at time *t* is given as *ϵ N*. Thus the probability *p*_not_(*t* + *dt*), that no pandemic has been initiated by time *t* + d*t* with d*t* being a small time step, can be expressed as

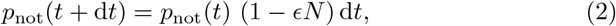

i.e., as the probability that no pandemic seeding event happened until time *t* times the probability that also no seeding event happened between *t* and *t* + d*t*. Therefore, the probability *p*_not_ decreases with time as

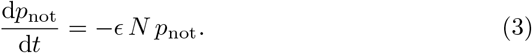

We obtain the probability that at least one such event has occurred (i.e. that a pandemic has been caused by time *t*) as *p* = 1− *p*_not_. Integrating Eq. (3) then leads to the pandemic risk of a single variant,

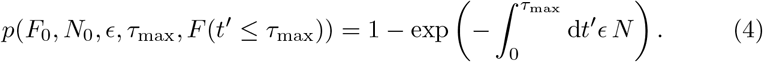

The above formula for pandemic risk is used for determinstic time-variations of fitness, where each outbreak of the same variant leads to the same growth curve in the infection center. For our pandemic risk measure we calculate the probability that a pandemic is caused by a variant within the specified global response time *τ*_max_ after its initial emergence.

The average pandemic risk *p*_pan_ of a viral type is finally determined by averaging the pandemic probability *p* across viruses from that type, with the initial variant fitness drawn from the characteristic distribution *f*_*F*_ (*F*_0_| *F*_m_, *σ*_*F*_). We calculate this average pandemic risk as

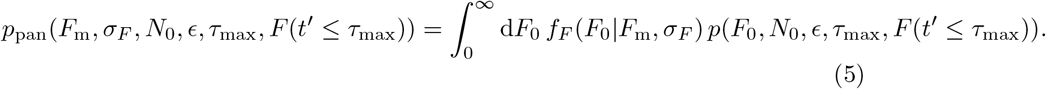

The integration starts at *F*_0_ = 0 (equivalent to *R* = 1), since with an initially negative fitness and small infection number *N*_0_, the virus will go extinct quickly due to the discrete nature of the underlying system, which we do not treat explicitly in our equations.

The single-variant risk *p*(*F*_0_ = *F*_m_) from Eq. (4), which describes the pandemic risk of a viral type with a single variant and therefore with no variance,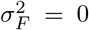, can be analytically expressed for the scenarios 1 and 2 considered below. The mean *p*_pan_(*F*_m_, *σ*_*F*_, *N*_0_, *ϵ, τ*_max_, *F* (*t′* ≤*τ*_max_)) (Eq. (5)) can be calculated numerically for those scenarios with deterministic fitness variation. We calculate this mean pandemic risk for various parameter combinations and we investigate and interpret its dependence on the different model parameters.

With the zoonotic shift rate we can, from *p*_pan_, calculate the expected frequency, at which pandemic variants of a certain type occur, as *µ*_shift_ *p*_pan_.

In the following, we first consider the pandemic risk directly for two different scenarios of the time-variation of fitness during the initial outbreak in the infection center: with time-invariant fitness and with linearly decreasing fitness. For each case we study the mean pandemic risk *p*_pan_ and analyze its dependence on the model parameters. We further consider a third scenario, with randomly varying fitness around a constant mean. For this last scenario we cannot directly obtain the average pandemic risk with an analytical formula, but in all cases we can determine a characteristic inflection time, beyond which international interventions will on average only have marginal effects.

## 3. Results

In order to determine pandemic risk under different outbreak conditions, we consider different temporal variations of fitness in the infection center. We assume the changes of disease fitness during the initial outbreak to mainly be caused by changes within the host population and the environment, and not due to mutation of the virus. For that we assume that the mutation rate of the virus is sufficiently small that it does not evolve much during the initial disease outbreak, which we focus on here. This situation includes SARS-CoV-2, where mutants only started playing an important role several months after the initial outbreak, but it does not include very highly mutable viruses like HIV that evolves significantly even within individual hosts. If the disease causes mild symptoms and only few hospitalizations, it likely stays unnoticed for an extended period and the disease growth rate can stay constant in a large infection center, where the supply of susceptible individuals stays high and no non-pharmaceutical interventions are put in place within *τ*_max_. This is the first scenario that we analyze; i.e., a constant fitness with time.

For the second scenario we assume that, due to hospitalizations, prevalent symptomatic infection or due to regular monitoring of zoonotic shifts, the disease is detected and early human interventions are put in place within the infection center. These can include the wearing of personal protective equipment, social distancing, reduced gatherings, or a combination of the above. Such interventions reduce the contact rate between infected and susceptible people and therefore the fitness. Fitness can also decrease due to the reduction of susceptible people in the infection center. As a simple example of a gradual fitness reduction we consider a linear decrease in fitness with time.

The third scenario is random variation of fitness, e.g. through fluctuating human behavior and non-sustained interventions. Such random variations of disease growth rates are represented by a Gaussian noise around a mean fitness.

Real disease outbreaks show a wide range of temporal variations, many of which can be characterized by combinations of the three scenarios we consider. For example, Thompson et al. [20] found that for different outbreaks of Ebola, pandemic flu, and MERS, the reproduction number varied in different fashions, being either roughly constant, decreasing or fluctuating.

### Scenario 1: Constant fitness

We first consider the scenario in which the fitness stays constant within the time scale *τ*_max_. In this case the number of infected people in a large infection center grows on average exponentially, following Eq. (1), as

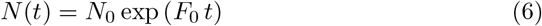

The pandemic risk for one variant with fitness *F*_0_ is then calculated using Eqs. (4) and (6) as

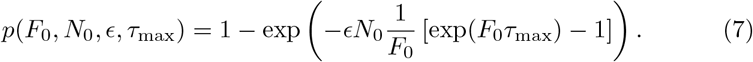

We numerically calculate the mean pandemic risk of the corresponding viral type with mean fitness *F*_m_ and fitness variance 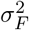 using Eq. (5) for a range of parameter values (Figs. 2 and 3). We compare this numerical result with the analytical zero-variance risk from Eq. (7) with *F*_0_ = *F*_m_.

**Figure 2.**
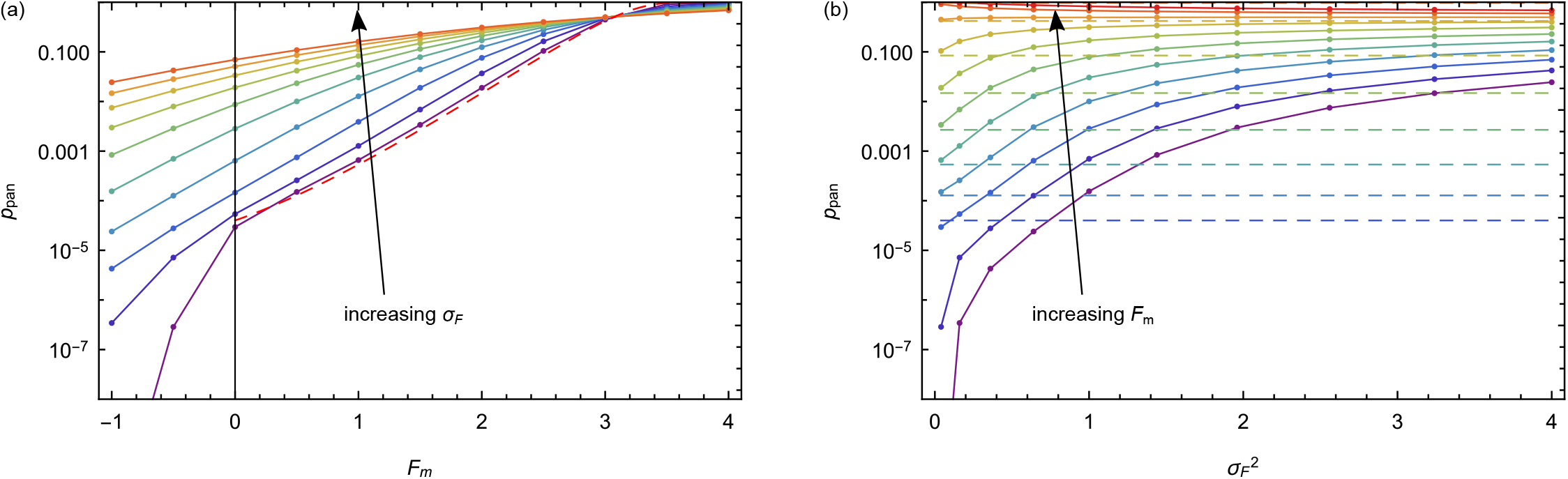
Mean pandemic risk of a viral type as function of mean fitness *F*_m_ (a) and of fitness variance 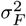 (b) for time-independent fitness. Colors from purple to orange indicate increasing values of *σ*_*F*_ from 0.2 to 2 in steps of 0.2 (a) and of *F*_m_ from -1 to 4 in steps of 0.5 (b). Circles joined by solid lines indicate numerical calculations using Eqs. (5) and (7); dashed lines represent the respective zero-variance risks from Eq. (7) with *F*_0_ = *F*_m_. The values of the parameters that are not varied in the respective graphs are *N*_0_ = 1, *ϵ* = 10^*−*5^ and *τ*_max_ = 4.

**Figure 3.**
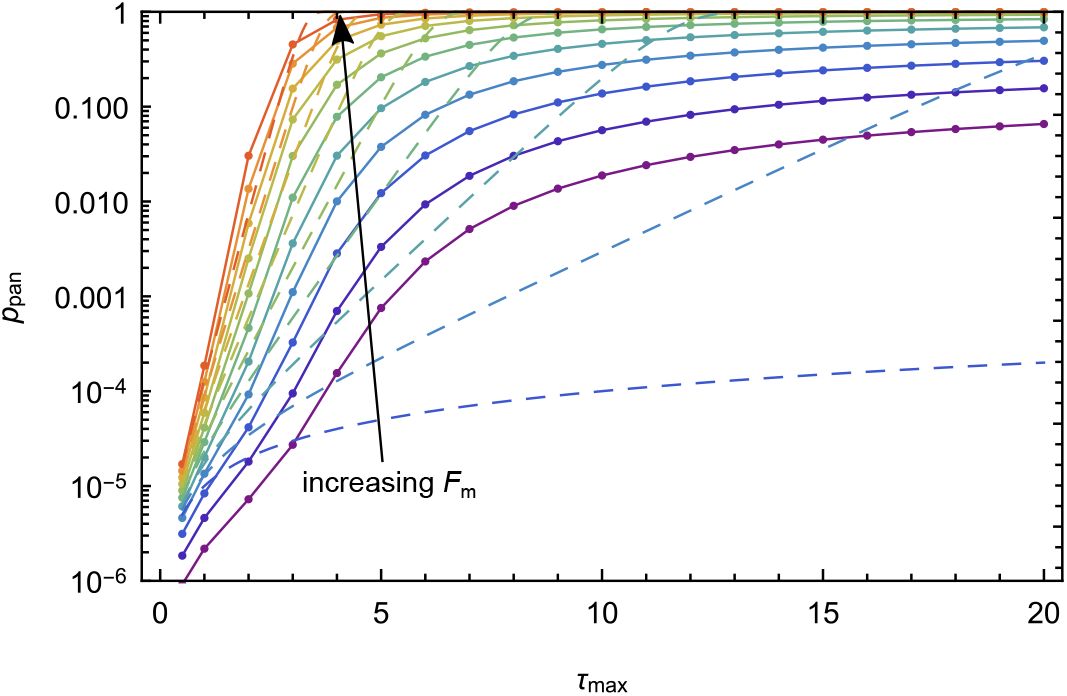
Mean pandemic risk of a viral type at various times *τ*_max_ for time-independent fitness. Colors from purple to orange indicate increasing values of mean fitness *F*_m_ from -1 to 4 in steps of 0.5. Circles joined by solid lines indicate numerical calculations using Eqs. (5) and (7); dashed lines represent the respective zero-variance risks from Eq. (7) with *F*_0_ = *F*_m_. The values of the parameters that are not varied in the graph are 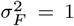, *N*_0_ = 1, and *ϵ* = 10^*−*5^.

For this single-variant risk (Eq. (7)) we expect that the pandemic risk first increases almost exponentially with fitness, before asymptotically approaching 1. This can be seen in Fig. 2a, which shows a nearly linear increase of the logarithm of pandemic risk with mean fitness for most of the observed fitness range before the asymptotic approach to 1.The single-variant risk agrees with the mean pandemic risk only for very low variance, while the mean risk can be

up to several orders of magnitude higher than the single-variant result in the case of wide fitness distributions, as long as *F*_m_ *>* 0 and if *F*_0_ is not too large. In this case, the pandemic risk initially increases with the variance 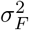 in a roughly exponential fashion, before tapering and asymptotically approaching 1 (Fig. 2b). This risk-enhancing effect of fitness variance in a symmetric distribution, which might seem surprising, mainly happens due to the averaging of a convex function as long as the pandemic risk is low. In this case *p*(*F*_0_) is convex and therefore

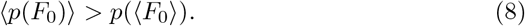

So, even if two strains are equally higher or lower in fitness than the mean, and so their average fitness equals *F*_m_, the contribution to the average of *p* from the higher-fitness strain more than makes up for that from the low-fitness strain.

A virus family, which is adapted to a reservoir host that is genetically very distantly related to us, but sometimes comes in physical proximity to humans or animals that humans interact with, can lead to many unsuccessful spillovers with on average very low fitness. But with a wide fitness distribution it can by chance sometimes produce fit variants and such rare events can result in pandemics. For example, a virus type with mean fitness *F*_m_ = 0.5 (equivalent to reproduction number *R* = 1.5) and with variance 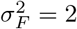, as well as *N*_0_ = 1, *ϵ* = 10^*−*5^, *τ*_max_ = 4 has a mean pandemic risk of around 4%, which is a hundred times higher risk compared to the case of a zero-variance fitness distribution.

When the variant fitness *F*_0_ is high, the single-variant risk (Eq. (7)) passes an inflection point 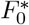, at which 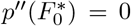. Thus, the convexity condition (Eq. 8), which holds for types with lower fitness where curvature is positive (*p ″* (*F*_0_) *>* 0) reverses for higher fitness values and we see in Fig. 2a that above the inflection point (here when 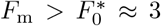), i.e. when *p ″* (*F*_0_) *<* 0, wide distributions with high variance have a reduced mean pandemic risk compared to the zero-variance case. At high mean fitness, those fitness variants, which have even higher fitness than the mean, cause a diminishing return in terms of pandemic risk since the pandemic probability is restricted by the upper limit of 1, while the co-occurring low-fitness variants cause the mean risk to decrease.

Virus types with negative mean fitness *F*_m_ *<* 0 and small variance have very low pandemic risk, since most variants produced by such a type will have negative growth rates. The pandemic risk reduces rapidly due to the diminishing portion of the fitness distribution that is above the extinction threshold *F*_0_ = 0, which for specific *F*_m_ and *σ*_*F*_ is given as

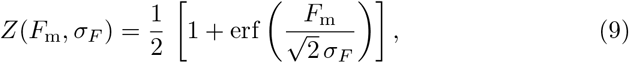

and which approaches 0 for negative mean fitness when *F*_m_ « −*σ*_*F*_.

As function of response time (Fig. 3), the mean pandemic risk again initially increases roughly exponentially for short response times before tapering towards an upper value, which is given by the proportion *Z*(*F*_m_, *σ*_*F*_) of positive-fitness strains (Eq. (9)). For the parameter values we consider, if the global response is delayed by only few disease generations, the pandemic risk can increase over several orders of magnitude. For example, if the infection period is around 1 week, the pandemic risk of a viral type with *F*_m_ = 1, 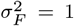, *N*_0_ = 1 and *ϵ* = 10^*−*5^ can increase from around 1/500 of a percent after the first week to more than 30% after a two-months delay (after 8 disease generations).

With finite fitness variance the increase with *τ*_max_ is initially larger than for zero-variance (Eq. (7)), due to the convex nonlinearity as function of *F*_0_ at short times. However, when global response time is slow, at long *τ*_max_, the risk starts levelling off towards its upper limit after passing the inflection point, where *p ″* (*F*_0_) changes sign. At long times, again, a large variance 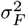 becomes disadvantageous for the virus due to the diminishing returns noted above. This means that the mean pandemic risk at long times is larger if the fitness of the newly emerging strains in a virus family exhibit no variance from a single value.

This zero-variance risk approaches its upper limit in the long-time limit rapidly with a double exponential function 1 — *p*(*τ*_max_) ∼ 1*/* exp(*λ* exp(*F*_m_*τ*_max_)) of time (Eq. (7)). The pandemic risk for finite variance, on the other hand, approaches its asymptotic limit much slower with 1 — *p*_pan_(*τ*_max_) ∼ 1*/* exp(*λτ*_max_)) with a single exponential. The increase of mean pandemic risk at long times is dominated by the lowest-fitness variants at the extinction threshold, with *F*_0_ →0, which have not had the chance yet to seed a pandemic, and which get there only at a low rate.

The effect of fitness variance leading to reduced risk at long times or at high mean fitness can only decrease risk down to a certain point, and not by orders of magnitude as opposed to the contrasting risk-enhancing effect of wide distributions, which applies before the inflection point of *p*(*F*_0_). We can understand this with the following intuitive argument. The pandemic risk of a variant can be expressed as *p*(*F*_0_) = 1 − exp(−*f* (*F*_0_)), where *f* (*F*_0_) = *ϵ* [*N* (*F*_0_) − *N*_0_]*/F*_0_ is a convex function of *F*_0_ (see Eq. (7)). As long as *F*_0_ and *τ*_max_ are small such that *f <* 1, the pandemic risk can be approximated with the convex function *p*(*F*_0_) ≈ *f* (*F*_0_) (using a first order Taylor approximation around *f* = 0). This means that the inflection point (separating convex from concave curvature) is around the point, where 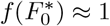, and at this point the pandemic risk 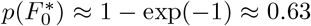 is already quite high. From this we conclude that the risk-reducing effect of wide distributions beyond the inflection point is only marginal because it only takes effect when the risk is already high. So, virus families characterized by a wide variance in the fitness of emergent variants that can infect humans generally pose a larger pandemic risk.

With increasing response time *τ*_max_, the pandemic risk not only passes an inflection point with respect to *F*_0_, but also an inflection time *t*^*∗*^, where *p″* (*t*^*∗*^) = 0, i.e. *p″* (*τ*_max_) changes sign from positive to negative. This is again around the point, where the pandemic risk *p*(*τ*_max_) = 1— exp(−*g*(*τ*_max_)) ≈1 −exp(− 1) ≈ 0.63 approaches its upper limit, i.e., when pandemic risk is no longer approximated by the convex (exponential) function *g*(*τ*_max_) = *ϵ* [*N* (*F*_0_) −*N*_0_]*/F*_0_. This means that changes in global response time matter significantly, due to exponentially increasing risk with time, before the inflection time, when *τ*_max_ *< t*^*∗*^. At longer times, *τ*_max_ *> t*^*∗*^, however, due to a diminishing return, varying the response time has an only marginal effect on pandemic risk, which then is already high and close to its upper limit. As illustrated above, global intervention can at this late point only prevent pandemic seeding of the least fit strains of the respective type. These findings indicate that overall a virus family is likely to be most dangerous, if it can seed an outbreak by causing a disease that is such that it takes long for humans to realize that a new virus is circulating.

The inflection time can be calculated by differentiating Eq. (3) with respect to time to obtain

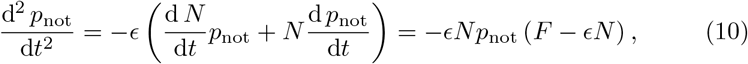

where we used the infection dynamics equation

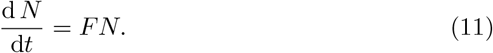

Therefore the inflection time is in general determined by

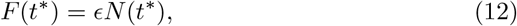

which leads in this case of constant fitness to

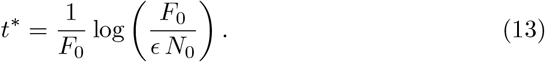

For an example virus with an infection period of one week, and with *F*_0_ = 1, *ϵ* = 10^*−*5^, *N*_0_ = 1, the dimensional inflection time would be around 3 months (12 infection periods), after which a global response could be classified as too late.

### Scenario 2: Decreasing fitness

As an alternative scenario we consider a disease which is such that the fitness of the emergent viral strain decreases with time. This could happen either because it is quickly detected in the infection center and early control measures and behavioral changes are put in place, or because the fraction of susceptible people in the infection center decreases notably. Here we consider a simple case of a linearly decreasing fitness *F* (*t*) = *F*_0_(1 − *t/τ*_*F*_) declining from an initial fitness *F*_0_ ∼*f*_*F*_ (*F*_0_ |*F*_m_, *σ*_*F*_). Here *τ*_*F*_ is an additional parameter representing the typical time scale, within which the local interventions or other mechanisms that lead to the fitness decrease fully take effect. The mean number of infected individuals in this case is calculated using Eq. (1) as

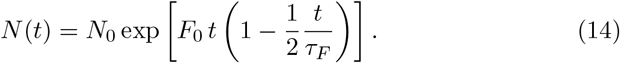

The pandemic probability for a single variant with initial fitness *F*_0_ is calculated with Eqs. (4) and (14) as

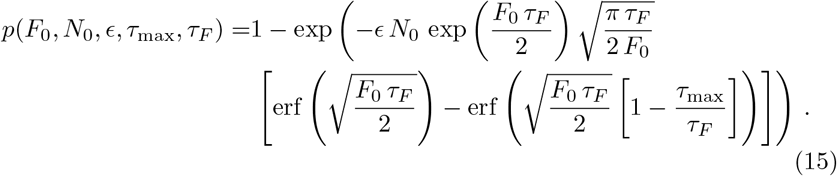

The above formula is valid for *τ*_max_ ≤ 2*τ*_*F*_. For longer response times the risk will stay at *p*(*τ*_max_ *>* 2*τ*_*F*_) = *p*(*τ*_max_ = 2*τ*_*F*_) ≈*ϵN*_0_*τ*_*F*_, since in that case the disease in the infection center will have reached an extinction threshold when the time equals 2*τ*_*F*_. By then the infection number will on average have decreased to the initial number *N*_0_ *>* 0, and assuming a small *N*_0_ plus the now negative and further decreasing fitness the disease will stochastically reach extinction soon after. From the single-variant risk we again obtain the mean pandemic risk numerically using Eq. (5), where we average over *F*_0_ ∼*f*_*F*_ (*F*_0_ |*F*_m_, *σ*_*F*_).

With such a linearly decreasing fitness the cumulative number of infections is given by:

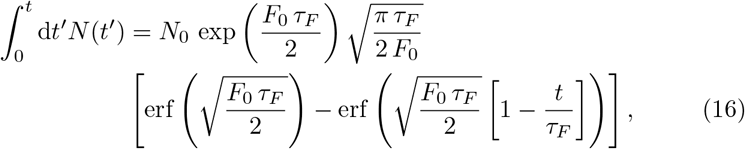

This quantity increases with time following an error function. Such a functional dependence is indeed what was observed for the cumulative number of infected people in various countries that implemented a wide range of social-distancing policies during the COVID-19 pandemic [21].

Both the zero-variance risk (Eq. 15) and the averaged pandemic risk increase with *τ*_*F*_ at first before *τ*_*F*_ ≫ *τ*_max_, after which they asymptotically approach the pandemic risk for constant *F* (*t*) = *F*_0_ (Fig. 4a). This shows that local responses, which often are more direct and faster in implementation than a global response (with *τ*_F_ *< τ*_max_) can reduce pandemic risk by several orders of magnitude via early suppression of disease spread in the infection center.

**Figure 4.**
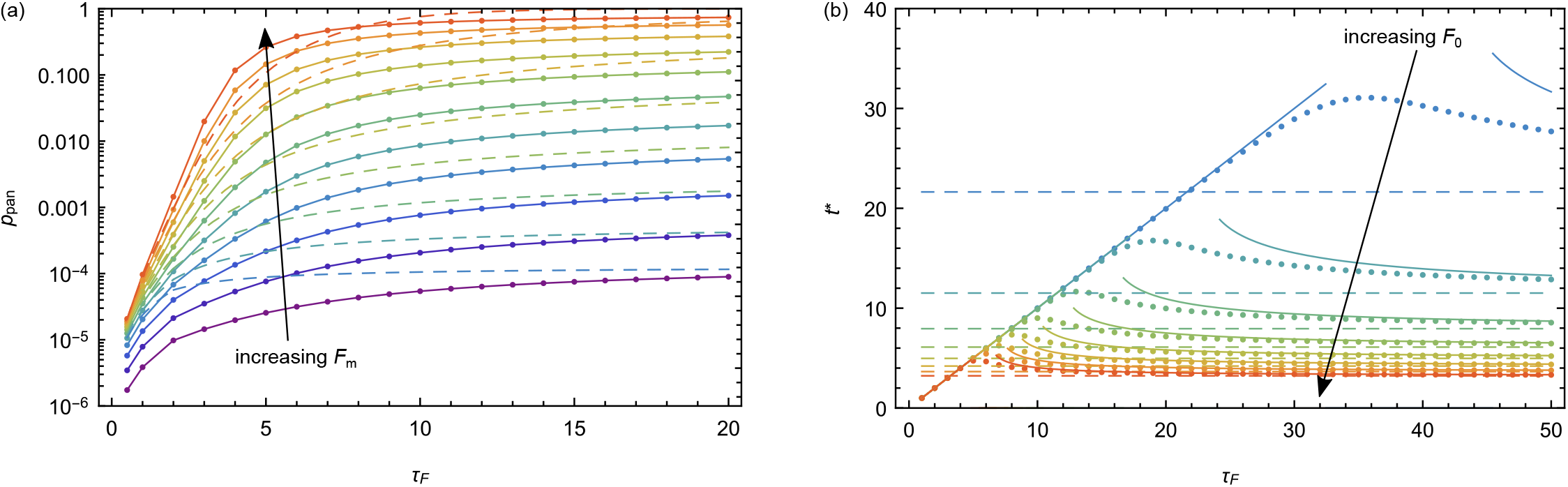
Mean pandemic risk of a viral type (a) and pandemic inflection time for a viral variant (b) in the case of linearly decreasing fitness, shown as function of the fitness decline time scale *τ*_*F*_. Colors from purple to orange indicate increasing values of mean fitness *F*_m_ from -1 to 4 in steps of 0.5 (a) and correspondingly, colors from blue to orange indicate increasing values of fitness *F*_0_ from 0.5 to 4 in steps of 0.5 (b). (a) Circles joined by solid lines indicate numerical calculations of mean risk using Eqs. (5) and (15); dashed lines represent the respective zero-variance risks from Eq. (15) with *F*_0_ = *F*_m_. (b) Circles represent numerical calculations of *t*^*∗*^ (Eq. (17)); solid lines represent the analytical approximations Eq. (18) and Eq. (20) for small and large *τ*_*F*_, respectively, dashed lines represent the inflection time for constant fitness *F* (*t*) = *F*_0_ (Eq. (13)). The values of the parameters that are not varied in the respective graphs are *σ*_*F*_ = 1, *N*_0_ = 1, *ϵ* = 10^*−*5^ and *τ*_max_ = 4.

The inflection time for the case of decreasing fitness is again calculated with Eq. (12) and is determined by

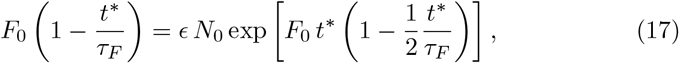

which we calculated numerically as function of *τ*_*F*_ and *F*_0_ (Fig. 4b). At small *τ*_*F*_ the inflection time increases with *τ*_*F*_ as

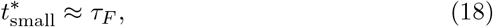

since then the inflection point is determined by the turning point (maximum) of the infected population in the infection center.On the other hand, when *τ*_*F*_→ ∞ the inflection time decreases again and approaches the inflection time 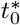 for constant fitness *F*_0_ (Eq. (13)). The decrease of the inflection time at long *τ*_*F*_ can be approximated as

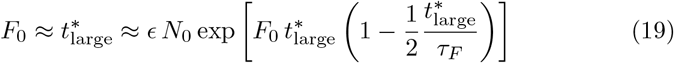

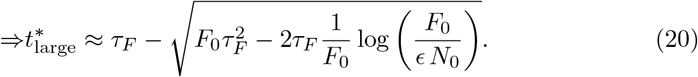

If the inflection time marks the same high pandemic risk in all cases, it would always be better to have a long inflection time, for pandemic risk control. For long intervention times *τ*_*F*_, where the upper limit of pandemic risk is roughly constant, this is the case and long inflection times then indicate a human advantage. However, for short local intervention times *τ*_*F*_ the upper limit of pandemic risk decreases with decreasing *τ*_*F*_, which is due to the effective control and extinction of the local outbreak. This shows the different effects of local interventions: more rapid interventions either decrease the upper limit of pandemic risk (at short *τ*_*F*_) or delay the time at which a high pandemic risk is reached (at long *τ*_*F*_) thereby giving more time for global interventions.

### Scenario 3: Randomly varying fitness

In addition to systematic changes of fitness, there can be random variations due to uncoordinated, non-sustained interventions or other human behavioral fluctuations. Here, as example of a random fitness variation in time, we consider fitness that fluctuates around a deterministic mean *F*_0_ as *F* (*t*) = *F*_0_ + *F*_r_*ξ*(*t*) with a Gaussian random variable *ξ*(*t*) and the characteristic amplitude *F*_r_ of the fluctuations. In this case we calculate the mean number of infections in the infection center (see Appendix A for details), averaged over different random fitness trajectories as

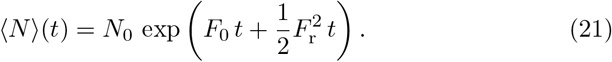

Although there is no simple formula in this case for the pandemic risk, averaged over random trajectories, we can calculate the average inflection time ⟨ *t*^*\**^⟩ as a function of the fitness parameters *F*_0_ and *F*_r_ using Eqs (12), (21) and the mean fitness ⟨ *F*⟩ (*t*) = *F*_0_ across different disease outbreaks of the same variant. We obtain

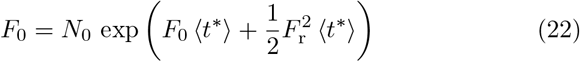

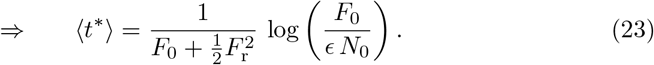

The inflection time again marks the time point, after which a global response can on average only lead to a marginal reduction of risk. Such a late response would in most individual outbreak situations be too late to prevent a global outbreak.

The inflection time decreases with increasing fitness components *F*_0_ and *F*_r_ (Fig. 5). For a virus type, whose different strains have baseline fitness *F*_0_ ∼ *f*_*F*_ (*F*_0_|*F*_m_, *σ*_*F*_) distributed around a mean *F*_m_, a delay in global response with *τ*_max_ *>*⟨ *t*^*\**^ ⟩ signifies that the response can only affect those strains significantly which on average grow slower than the mean, with *F*_0_ *< F*_m_, and it would have little effect on the majority of strains. For *F*_m_ = 1 and assuming a relatively large fluctuation amplitude *F*_r_ = 2, and *ϵ* = 10^*−*5^, this characteristic inflection time is⟨ *t*^*\**^ ⟩ (*F*_m_, *F*_r_) = 4 infection periods, while without random fluctuations in time, *F*_r_ = 0, it would be three times longer with *t*^*\**^(*F*_m_, *F*_r_) = 12 infection periods, thus on average giving more time to react in the case of low fitness fluctuations.

**Figure 5.**
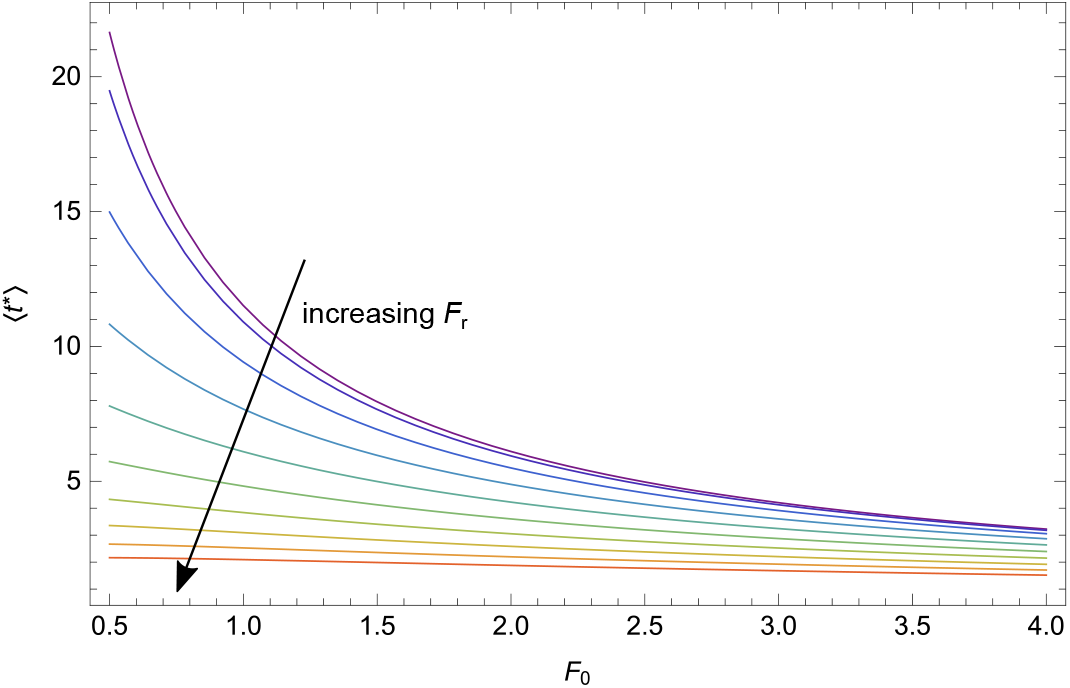
Inflection time as function of baseline fitness *F*_0_ (Eq. (23)) in the case of randomly varying fitness. Colors from purple to orange indicate increasing values of the amplitude of fitness fluctuations in time, *F*_r_, from 0 to 3 in steps of 0.3. The values of the parameters that are not varied in the graph are *N*_0_ = 1 and ϵ = 10−5.

## 4. Discussion

With a minimal pandemic risk model we have investigated how pandemics emerge from characteristically different viral types. We analyzed how pandemic risk and frequency are influenced by key viral and human population traits: mean and variance of the disease growth rate or fitness, zoonotic shift frequency, world transmission rate, local and global response times and variations in fitness with time.

We show that wide fitness distributions can drastically increase pandemic risk compared to narrow distributions around the same mean fitness, because the functional dependence of pandemic risk on viral fitness is initially convex. We further identify an inflection point in pandemic risk variation with respect to fitness, beyond which the convexity condition is not valid and narrow fitness distributions become more dangerous compared to wide distributions in terms of pandemic risk.

It is well known that swift collaborative action is important to control the global spread of novel pathogens [22]. But how rapid do these interventions need to be, i.e. what is the relevant time scale? We identify this time scale to be the pandemic inflection time, after which human responses are likely too late to stop the world-wide spread of the disease. Previous studies have discussed an epidemic inflection time [23, 24], which corresponds to the time point at which the number of infected people in a population reaches its maximum (i.e. its turning point). This time scale relies on the leveling off of the cumulative number 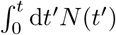 of infections within a relatively short time compared to the global disease transmission time scale. It therefore represents a special case of our pandemic inflection time. The epidemic inflection time coincides with our measure only in scenario 2, in the case of rapid and complete control (with small *τ*_*F*_) of the local outbreak. The pandemic inflection time, however, describes the time point, at which the pandemic risk starts levelling off in general. This can be due to a local turning point as discussed in the context of scenario 2, but it can also simply occur due to the approach of the upper limit of pandemic probability (100%), at which point most outbreaks would have already caused a pandemic and only few strains, at the low-fitness end of the distribution, would not yet have reached their final (pandemic) outcome.

Local interventions, which gradually reduce the disease growth rate (scenario 2), can by themselves prevent pandemics only if they are able to rapidly extinguish the outbreak. This is what happened during the Ebola outbreaks, and to some extent for MERS. Both these diseases were caused by highly lethal viruses, and thus were quickly noticed and controlled locally. If, however, local interventions reduce the spread in the infection center slowly and incompletely, this merely postpones the inflection time point. In the latter case global interventions are needed, but are given more time to be put into action. Linear fitness decrease in the infection center due to such interventions leads to an error function-type increase of cumulative cases. This characteristic time-dependent behavior of cumulative case numbers has recently been observed in several countries, where different social-distancing measures against COVID-19 were put in place [21]. With the model of linearly decreasing fitness we thus provide one mechanistic explanation for such trends.

As long as the disease is not completely eradicated and immunization measures are not yet developed, it is important that control measures are consistently applied to systematically decrease fitness in the infection center instead of relying on intermittent interventions and natural behavioral adaptations, which might lead to large fitness fluctuations in time. Such random fluctuations are demonstrated to on average shorten the inflection time and therefore give global interventions less time to effectively prevent a global outbreak (scenario 3).

The richness and diversity of infectious disease outbreaks have increased since 1980 [3], which is likely due to human factors such as the continuous increases in population size, density and connectedness as well as human occupation of new locales [25, 26]. Several of our model parameters can be influenced by human population trends in ways that can explain a rising pandemic risk. The zoonotic shift rate and fitness variance across strains have increased from an increased frequency of animal-human contact due to increases in livestock and wildlife trade and human encroachment in new areas (via deforestation and agricultural expansion) [15, 6]. The mean fitness of human variants could have increased due to increased human population density, especially in urban environments, which increases the contact rate between humans and therefore the disease growth rate. Finally, the world transmission rate is undoubtedly higher due to increased global connectedness and travel.

On the other hand, a decrease in disease fitness and world transmission rate, as well as increased speeds of response (local ∼ 1*/τ*_*F*_ or global ∼ 1*/τ*_max_) are achieved through technological and scientific advances leading to improved surveillance as well as through informed and swiftly implemented policies. Such concerted efforts decrease pandemic risk; here it appears that global mobilization issues present a greater source of delayed response than surveillance capacity [22].

In the face of sparse data on newly emerging infectious diseases [14] and to make predictions for diverse viral pathogens, the simplicity and generality of our model are a strength. Representing viral types with simple fitness distributions of new variants, without resolving specific evolutionary paths and fitness landscape topologies, makes our model analytically tractable while still capturing important stochastic effects, and could potentially provide mechanistic underpinnings to empirical observations or more detailed descriptions. In particular, we hope that the importance of the inflection points revealed by our analyses will help guide data-driven public policy.

## Data Availability

all information is included in the manuscript.

## Acknowledgements

This research was supported by the Ragon Institute of MGH, MIT, and Harvard and NIH grant # R01HL120724-01A1. MK acknowledges support by the NSF through grants # DMR-1708280 and # PHY-2026995.

## Author contributions

AKC instigated the project. JD, MK and AKC designed research, analyzed the results and wrote the paper. JD performed the calculations and generated all figures.

## Appendix A. Calculation details for randomly varying fitness (scenario 3)

In the case of random fitness fluctuations we have assumed

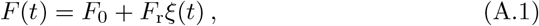

with a time-independent component *F*_0_, and Gaussian fluctuations of the fitness around *F*_0_ with typical amplitude *F*_r_. The random variable *ξ*(*t*) is a Gaussian white noise characterized by a unit variance, ⟨ *ξ*(*t*)*ξ*(*t′*) ⟩ = *δ*(*t − t′*), and zero mean, ⟨ *ξ*(*t*) ⟩ = 0.

Accordingly, the infected population size for a certain variant spreading in one population varies randomly in time, i.e.,

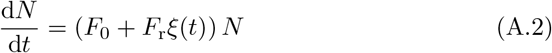

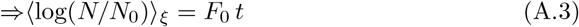

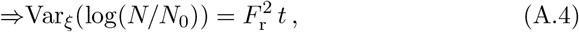

with the mean and variance (subscript *ξ*) as summary statistics across various random fitness trajectories (averaging over different outbreaks of disease caused by the same variant). The mean number of infections after time *t* is then calculated as

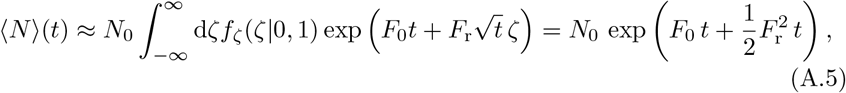

where the random variable *ζ* is also Gaussian following the normal distribution *f*_*ζ*_(*ζ*|0, 1) with zero mean and unit variance.

## References

[1] W. Qiu, S. Rutherford, A. Mao, C. Chu, The Pandemic and its Impacts, Health, Culture and Society 9–10 (2016–2017) 1–11. doi:10.5195/hcs.2017.221.

[2] K. E. Jones, N. G. Patel, M. A. Levy, A. Storeygard, D. Balk, J. L. Gittleman, P. Daszak, Global trends in emerging infectious diseases, Nature Letters 451 (2008) 990–993. doi:10.1038/nature06536.

[3] K. F. Smith, M. Goldberg, S. Rosenthal, L. Carlson, J. Chen, C. Chen, S. Ramachandran, Global rise in human infectious disease outbreaks, Journal of the Royal Society Interface 11 (20140950) (2014). doi:10.1098/rsif.2014.0950.

[4] J. A. Oldekop, R. Horner, D. Hulme, R. Adhikari, B. Agarwal, M. Alford, O. Bakewell, N. Banks, S. Barrientos, T. Bastia, et al., COVID-19 and the case for global development, World Development 134 (105044) (2020). doi:10.1016/j.worlddev.2020.105044.

[5] R. Baldwin, B. Weder di Mauro, Economics in the Time of COVID-19, CEPR Press, 2020.

[6] C. R. Parrish, E. C. Holmes, D. M. Morens, E.-C. Park, D. S. Burke, C. H. Calisher, C. A. Laughlin, L. J. Saif, P. Daszak, Cross-species virus transmission and the emergence of new epidemic diseases, Microbiology and Molecular Biology Reviews 72 (3) (2008) 457–470. doi:10.1128/MMBR.00004-08.

[7] R. K. Plowright, C. R. Parrish, H. McCallum, P. J. Hudson, A. I. Ko, A. L. Graham, J. O. Lloyd-Smith, Pathways to zoonotic spillover, Nature Reviews Microbiology 15 (2017) 502–510. doi:10.1038/nrmicro.2017.45.

[8] A. K. Chakraborty, A. S. Shaw, Viruses, Pandemics, and Immunity, MIT Press, 2020.

[9] R. N. Thompson, C. A. Gilligan, N. J. Cunniffe, Will an outbreak exceed available resources for control? Estimating the risk from invading pathogens using practical definitions of a severe epidemic, Journal of the Royal Society Interface 17 (172) (2020) 1–13. doi:10.1098/rsif.2020.0690.

[10] F. Brauer, P. Van den Driessche, J. Wu, Mathematical Epidemiology, Springer, Berlin, Germany, 2008. doi:110.1007/978-3-540-78911-6.

[11] M. Lipsitch, W. Barclay, R. Raman, C. J. Russell, J. A. Belser, S. Cobey, P. M. Kasson, J. O. Lloyd-Smith, S. Maurer-Stroh, S. Riley, et al., Viral factors in influenza pandemic risk assessment, eLife 5 (2016) e18491. doi: 10.7554/eLife.18491.

[12] J. O. Lloyd-Smith, S. Funk, A. R. McLean, S. Riley, J. L. Wood, Nine challenges in modelling the emergence of novel pathogens, Epidemics 10 (2015) 35–39. doi:10.1016/j.epidem.2014.09.002.

[13] R. Antia, R. R. Regoes, J. C. Koella, C. T. Bergstrom, The role of evolution in the emergence of infectious diseases, Nature 426 (2003) 658–661. doi: 10.1038/nature02104.

[14] C. J. E. Metcalf, J. Lessler, Opportunities and challenges in modeling emerging infectious diseases, Science 357 (6347) (2017) 149–152. doi: 10.1126/science.aam8335.

[15] S. S. Morse, J. A. Mazet, M. Woolhouse, C. R. Parrish, D. Carroll, W. B. Karesh, C. Zambrana-Torrelio, W. I. Lipkin, P. Daszak, Prediction and prevention of the next pandemic zoonosis, The Lancet 380 (9857) (2012) 1956–1965. doi:10.1016/S0140-6736(12)61684-5.

[16] K. J. Olival, P. R. Hosseini, C. Zambrana-Torrelio, N. Ross, T. L. Bogich, P. Daszak, Host and viral traits predict zoonotic spillover from mammals, Nature 546 (2017) 646–650. doi:10.1038/nature22975.

[17] L. P. Shaw, A. D. Wang, D. Dylus, M. Meier, G. Pogacnik, C. Dessimoz, F. Balloux, The phylogenetic range of bacterial and viral pathogens of vertebrates, Molecular Ecology 29 (2020). doi:10.1111/mec.15463.

[18] J. S. Long, B. Mistry, S. M. Haslam, W. S. Barclay, Host and viral determinants of influenza A virus species specificity, Nature Reviews Microbiology 17 (2019) 67–81. doi:10.1038/s41579-018-0115-z.

[19] J. Gerges Harb, H. A. Noureldine, G. Chedid, M. N. Eldine, D. A. Abdallah, N. F. Chedid, W. Nour-Eldine, SARS, MERS and COVID-19: clinical manifestations and organ-system complications: a mini review, Pathogens and Disease 78 (2020) 1–7. doi:10.1093/femspd/ftaa033.

[20] R. N. Thompson, J. E. Stockwin, R. D. van Gaalen, J. A. Polonsky, Z. N. Kamvar, P. A. Demarsh, E. Dahlqwist, S. Li, E. Miguel, T. Jombart, et al., Improved inference of time-varying reproduction numbers during infectious disease outbreaks, Epidemics 29 (100356) (2019) 1–11. doi:10.1016/j.epidem.2019.100356.

[21] R. Marsland, P. Mehta, Data-driven modeling reveals a universal dynamic underlying the covid-19 pandemic under social distancing, medRxiv (2020). doi:10.1101/2020.04.21.20073890.

[22] S. J. Hoffman, S. L. Silverberg, Delays in Global Disease Outbreak Responses: Lessons from H1N1, Ebola, and Zika, American Journal of Public Health 108 (3) (2018) 329–333. doi:10.2105/AJPH.2017.304245.

[23] Z. Ma, Predicting the outbreak risks and inflection points of covid-19 pandemic with classic ecological theories, Advanced Science 7 (21) (2020) 2001530.

[24] Q. Duan, J. Wu, G. Wu, Y.-G. Wang, Predication of inflection point and outbreak size of covid-19 in new epicentres, arXiv:2007.07471 preprint (2020).

[25] R. A. Weiss, A. J. McMichael, Social and environmental risk factors in the emergence of infectious diseases, Nature Medicine 10 (12) (2004) S70–S76. doi:10.1038/nm1150.

[26] A. C. Ezeh, J. Bongaarts, B. Mberu, Global population trends and policy options, The Lancet 380 (2012) 142–148. doi:10.1016/S0140-6736(12)60696-5.

